# Real-world effectiveness of nirmatrelvir/ritonavir use for COVID-19: A population-based cohort study in Ontario, Canada

**DOI:** 10.1101/2022.11.03.22281881

**Authors:** KL Schwartz, J Wang, M Tadrous, BJ Langford, N Daneman, V Leung, T Gomes, L Friedman, P Daley, KA Brown

## Abstract

**Background:** Our objective was to evaluate the real world effectiveness of nirmatrelvir/ritonavir to prevent severe COVID-19 while Omicron and its subvariants predominate.

**Methods:** We conducted a population based cohort study in Ontario, Canada including all residents >17 years of age who tested positive for SARS-CoV-2 by PCR between 4 April and 31 August 2022. We compared nirmatrelvir/ritonavir treated patients to unexposed patients and measured the primary outcome of hospitalization or death from COVID-19, and a secondary outcome of death 1-30 days. We used weighted logistic regression to calculate weighted odds ratios (wOR) with 95% confidence intervals (CIs) using inverse probability of treatment weighting (IPTW) to control for confounding.

**Results:** The final cohort included 177,545 patients with 8,876 (5.0%) exposed and 168,669 (95.0%) unexposed individuals. The groups were well balanced with respect to demographic and clinical characteristics after applying stabilized IPTW. Hospitalization or death within 30 days was lower in the nirmatrelvir/ritonavir treated group compared to unexposed individuals (2.1% vs 3.7%, wOR 0.56; 95%CI, 0.47-0.67). In the secondary analysis, the relative odds of death was also significantly reduced (1.6% vs 3.3%, wOR 0.49; 95%CI, 0.39-0.62). The number needed to treat to prevent one case of severe COVID-19 was 62 (95%CI 43 to 80). Findings were similar across strata of age, DDIs, vaccination status, and comorbidities.

**Interpretation:** Nirmatrelvir/ritonavir was associated with significantly reduced risk of hospitalization and death from COVID-19 in this observational study, supporting ongoing use of this therapeutic to treat patients with mild COVID-19 at risk for severe disease.

## Introduction

Severe acute respiratory syndrome coronavirus 2 (SARS-CoV-2) infections have been associated with substantial morbidity and mortality and at times overwhelmed healthcare capacity in many jurisdictions since 2020. Immunization and immunity in those who have recovered from infection have substantially modified the disease course and improved outcomes from coronavirus disease 2019 (COVID-19). Emergent Omicron variants also have reduced virulence compared to the progenitor virus.(1) Antiviral therapies to treat milder infections and prevent severe outcomes such as hospitalization and death from COVID-19 are valuable additional tools in the global pandemic response. The EPIC-HR randomized controlled trial of nirmatrelvir/ritonavir identified an 89% reduction in progression to severe COVID-19 in treated participants at high risk of severe disease, compared to placebo. However, the trial was conducted between July and December 2021, prior to the emergence of the Omicron variant, and excluded vaccinated individuals as well as those with drug-drug interactions (DDIs).(2)

In real-world evaluations of nirmatrelvir/ritonavir while the Omicron variant of concern and subvariants are predominating, a significant protective effect was seen in adults 65 years of age and older in Israel, but not in adults 40-64 years.(3) A study from Hong Kong identified a significant protective effect in adults, albeit attenuated compared to the EPIC-HR trial.(4) Studies that have stratified by vaccination status have identified similar relative risk reductions in vaccinated cohorts, but with smaller absolute risk reductions due to the lower baseline risk of hospitalization or death from COVID-19.(3,5,6) All previous observational studies have risks of bias including residual confounding and immortal time bias.(7)

In Ontario, Canada nirmatrelvir/ritonavir became widely available, and universally funded for all patients, in the community by April 2022 with clinical criteria set by the government limiting access only to patients who were older, had comorbidities, and/or were undervaccinated.(8,9) The Ontario Science Table (OST) provided clinical practice guidance to Ontario clinicians on the use of therapeutics for COVID-19 with stricter high-risk criteria based on patients who were most likely to benefit from the limited supplies of antiviral drug at the time.(10) A large proportion of patients receiving nirmatrelvir/ritonavir in Ontario were expected to have been excluded from the EPIC-HR trial population, such as those previously vaccinated or receiving concomitant medications with significant DDIs. Observational data evaluating real-world use of this medication is critical to inform future policy and guidelines. Our objective was to evaluate the real world effectiveness of nirmatrelvir/ritonavir on health outcomes including hospitalization and death from COVID-19 while Omicron and its subvariants predominate.

## Methods

### Population and Setting

We conducted a population based cohort study in Ontario (Canada’s most populous province, with a population of 15 million in 2022). Ontario has universal health insurance which covers virtually the entire population. Nirmatrelvir/ritonavir was covered for all Ontarians irrespective of health insurance coverage. All Ontario residents between the ages of 18 and 110 years between 4 April 2022 and 31 August 2022 with a positive polymerase chain reaction (PCR) test for SARS-CoV-2 were assessed for study inclusion. Individuals were excluded if they were not Ontario residents or had invalid identifiers such as date of birth, or date of death was prior to test date. Hospitalized patients or nosocomial infections (i.e. patients hospitalized prior to or on the day of testing) were also excluded. The data are housed and were analyzed at ICES (formerly, The Institute for Clinical Evaluative Sciences) using unique encoded identifiers. ICES is an independent, non-profit research institute whose legal status under Ontario’s health information privacy law allows it to collect and analyze health care and demographic data, without consent, for health system evaluation and improvement. This study has Ethics Research Board approval by Public Health Ontario (2022-015.01).

### Study Design

We conducted an observational cohort study utilizing inverse probability of treatment weighting (IPTW) from propensity scores to adjust for confounding. A propensity score is defined as the probability of treatment assignment conditional on measured baseline covariates(11,12). IPTW weights individuals by the inverse probability of receiving nirmatrelvir/ritonavir in the exposed group and weights the unexposed group with the probability of receiving nirmatrelvir/ritonavir by creating a synthetic sample in which nirmatrelvir/ritonavir receipt is independent of measured baseline covariates. IPTW obtains unbiased estimates of average treatment effects (assuming no residual confounding).(11) Due to the very low propensity for receiving nirmatrelvir/ritonavir for some covariates, we used stabilized weights, which reduces the variability of the estimated treatment effect.(13) Variables included in the IPTW were selected *a priori* based on their clinically important risk of confounding and included age, sex, number of SARS-CoV-2 vaccine doses (0, 1, 2, or 3+), previous SARS-CoV-2 infection, time from last vaccine dose (14-89, 90-179, 180-269, or 270+ days), individual comorbidities (including chronic respiratory disease, chronic heart disease, hypertension, diabetes, immune compromised, autoimmune disease, dementia, chronic kidney disease, and advanced liver disease) (Appendix Table S1), long term care residence, and high versus standard risk as per the OST which is based on age, comorbidities, and number of vaccine doses received.(10)

### Data Sources and Definitions

Nirmatrelvir/ritonavir prescription data was obtained from the Ontario Drug Benefit (ODB) database which is >99% accurate in identifying outpatient prescription medications dispensed.(14) Nirmatrelvir/ritonavir was approved for use by Health Canada on 17 January 2022. Shortly thereafter, limited supplies were available from select COVID-19 assessment centres in Ontario for use; however these prescriptions were not captured in ODB. Beginning 4 April 2022, publicly-funded access to nirmatrelvir/ritonavir from community pharmacies became available for any Ontarian meeting the province’s eligibility criteria (Appendix Table S2). Dispensing through community pharmacies increased rapidly, reaching approximately 85% of all nirmatrelvir/ritonavir prescriptions during the study period. Therefore, to reduce the risk of misclassification bias, anyone testing positive for SARS-CoV-2 from one of 27 COVID-19 assessment centres dispensing nirmatrelvir/ritonavir were excluded from this study as their exposure status cannot be determined using our data (Figure 1). SARS-CoV-2 testing was obtained from the C19INTGR database which contains all SARS-CoV-2 PCR tests (but not antigen test results). Eligibility for PCR testing changed in December 2021 and was limited to specific groups including those eligible for treatment or healthcare workers. Vaccination status was obtained from the COVAXON database.(15) Comorbidity data was obtained from the Canadian Institute for Health Information (CIHI) databases, Ontario Health Insurance Plan (OHIP) database, as well as other validated disease specific cohorts at ICES (Appendix Table S1). The index date for exposed individuals was defined as the date nirmatrelvir/ritonavir was dispensed. A time-to-dispense (TTD) distribution was then created for the exposed cohort defined as the time in days from test positive to medication dispensing. To minimize immortal time bias a random index date was then assigned to the unexposed group based on the TTD distribution from exposed individuals. For example, 37% of the exposed group was dispensed nirmatrelvir/ritonavir on day 0; therefore if the random number generated for an unexposed individual was between 0 and 0.37, they were assigned a TTD=0 and their index date was defined as their test date (Appendix Figure S1). Any patient who died or was hospitalized for COVID-19 on, or prior to, their index date (dispense date for exposed individuals and simulated index date for unexposed) was excluded. DDI data was obtained from ODB but was limited to patients over 70 years of age for this study as ODB does not contain data on medications other than nirmatrelvir/ritonavir for younger patients. Potential DDI was defined as any severity level 1 or level 2 co-medications with an ODB claim with an overlap in days supplied and dispense date of nirmatrelvir/ritonavir, where level 1 included any co-medications contraindicated with nirmatrelvir/ritonavir (i.e. nirmatrelvir/ritonavir should not be prescribed as stopping the co-medication is insufficient to mitigate DDI) and level 2 included co-medications with clinically significant DDIs requiring a mitigation strategy while on nirmatrelvir/ritonavir (i.e. holding co-medication, dose/interval adjustment, use of alternative agent, management of side effects, additional monitoring) according to OST guidelines.(16) (Appendix Table S3) DDIs were not evaluated for patients <70 years of age.

**Figure 1:**
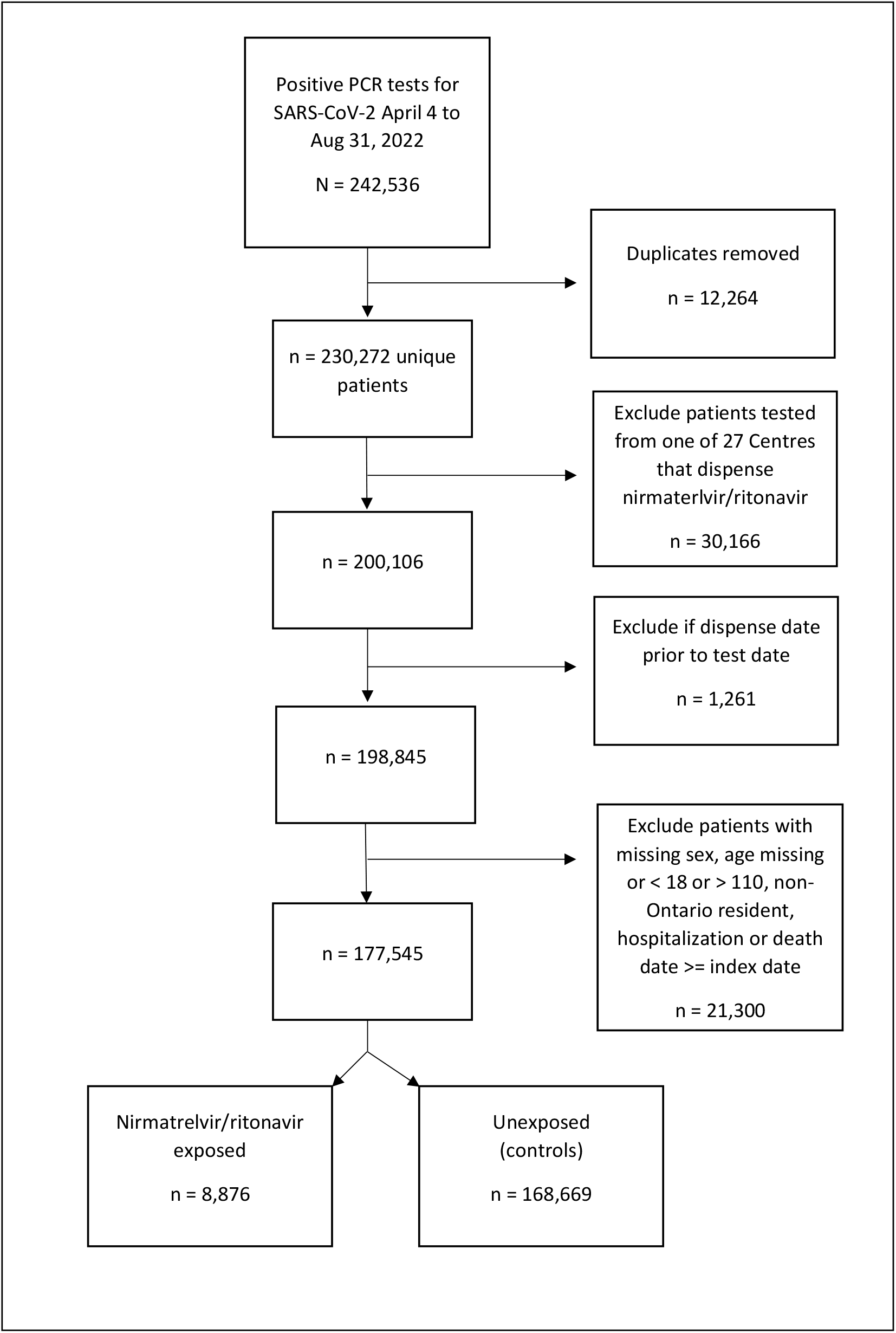
Cohort creation flow diagram

### Study Outcomes

The primary study outcome was the composite of hospitalization for COVID-19 or death 1-30 days after the index date. This was ascertained from the Case and Contact Management (CCM) database which is completed by local public health units for public health purposes and defines hospitalization as related to COVID-19 or not.(15) Secondary outcome was death 1-30 days from the index date. Preplanned subgroup analyses included stratification by age (≥70 or <70 years), vaccination status (0, 1-2, or 3+ doses), potential DDIs for those over 70 years of age (level 1,level 2 or no DDIs identified - Appendix Table S3), comorbidities (3 or more, or <3), long term care residents, OST high versus standard risk, and time (April-June 2022 or July to August 2022).

### Statistical Analysis

Unweighted and weighted (using stabilized weights) covariates were compared using standardized differences (SD), with SD<0.1 reflecting clinically insignificant differences. IPTW weighted exposures were evaluated for the outcomes using logistic regression models presented as weighted odds ratios (wOR) with 95% confidence intervals and p-values<0.05 representing statistical significance. Using weighted logistic regression models with calculated the number needed to treat (NNT) based on the absolute risk reductions with 95%CIs. Data were analyzed using SAS enterprise guide 9.4 (Cary, NC).

## Results

We identified 242,536 individuals who tested positive for SARS-CoV-2 by PCR between 4 April 2022 and 31 August 2022. After applying the study inclusion and exclusion criteria there were 8,876 individuals exposed to nirmatrelvir/ritonavir and 168,669 people who remained unexposed (Figure 1). Prior to weighting, the nirmatrelvir/ritonavir exposed cohort was predominately 70 years of age or older (72.5%), had received 3 or more vaccine doses (84.8%), had <3 comorbidities (57.1%), were standard risk by OST criteria (58.1%), and did not reside in long term care (68.5%). For those 70 years of age or older, 66.7% had 1 or more potential DDIs (Appendix Table S4). Prior to weighting, major between-group differences existed in almost all variables evaluated. Nirmatrelvir/ritonavir recipients were older, more likely to have 3+ vaccine doses, had more comorbidities, were more likely to meet OST high risk criteria, and reside in long term care. After weighting there were no clinically significant differences between any covariates represented by standardized differences <0.1 (Table 1 and Figure 2).

**Table 1:** Population characteristics pre- and post-applying stabilized weights with standardized differences (SD)

| variable | Unweighted population |  |  | Weighted population |  |  |
| --- | --- | --- | --- | --- | --- | --- |
|  | Exposed<br>n=8,876 | Unexposed<br>n=168,669 | Unweighted<br>SD | Exposed | Unexposed | Weighted<br>SD |
| **Age (years)** | 74.3 | 52.4 | 1.17 | 74.3 | 74.4 | 0.01 |
| **Female sex** | 59.3% | 63.4% | 0.08 | 59.3% | 59.4% | <0.01 |
| **Vaccine doses** |  |  |  |  |  |  |
| **0** | 5.3% | 6.2% | 0.04 | 5.3% | 5.3% | <0.01 |
| **1** | 1.0% | 1.0% | <0.01 | 1.0% | 1.0% | <0.01 |
| **2** | 9.0% | 17.0% | 0.24 | 9.0% | 9.1% | <0.01 |
| **3+** | 84.8% | 75.8% | 0.23 | 84.8% | 84.6% | <0.01 |
| **Previous SARS-CoV2 infection** | 4.6% | 6.9% | 0.10 | 4.6% | 4.6% | <0.01 |
| **Time from last vaccine dose** |  |  |  |  |  |  |
| **14-89 days** | 16.4% | 10.3% | 0.18 | 16.4% | 16.6% | 0.01 |
| **90-179 days** | 42.4% | 43.1% | 0.02 | 42.4% | 42.3% | <0.01 |
| **180-269 days** | 27.1% | 27.4% | 0.01 | 27.1% | 26.8% | 0.01 |
| **270+ days** | 14.2% | 19.2% | 0.13 | 14.2% | 14.4% | 0.01 |
| **Ontario Science Table risk group** |  |  |  |  |  |  |
| **High risk** | 41.9% | 15.1% | 0.62 | 41.9% | 42.5% | 0.01 |
| **Standard risk** | 58.1% | 84.9% | 0.62 | 58.1% | 57.5% | 0.01 |
| **Comorbidities** |  |  |  |  |  |  |
| **Chronic respiratory disease** | 35.2% | 24.2% | 0.24 | 35.2% | 35.7% | 0.01 |
| **Chronic heart disease** | 25.3% | 11.2% | 0.37 | 25.3% | 25.7% | 0.01 |
| **Diabetes** | 33.8% | 16.8% | 0.40 | 33.8% | 34.1% | 0.01 |
| **Immune compromised** | 15.9 | 6.0% | 0.32 | 15.9% | 16.8% | 0.02 |
| **Hypertension** | 68.4% | 32.3% | 0.77 | 68.4% | 68.7% | 0.01 |
| **Dementia** | 30.0% | 9.3% | 0.54 | 30.0% | 30.1% | <0.01 |
| **Autoimmune disease** | 13.0% | 5.0% | 0.28 | 13.0% | 14.1% | 0.03 |
| **Chronic Kidney Disease** | 12.5% | 5.9% | 0.23 | 12.5% | 12.6% | <0.01 |
| **Advanced Liver Disease** | 2.4% | 1.3% | 0.08 | 2.4% | 2.4% | <0.01 |
| **LTC resident** | 31.5% | 7.6% | 0.63 | 31.5% | 31.8% | 0.01 |

**Figure 2:**
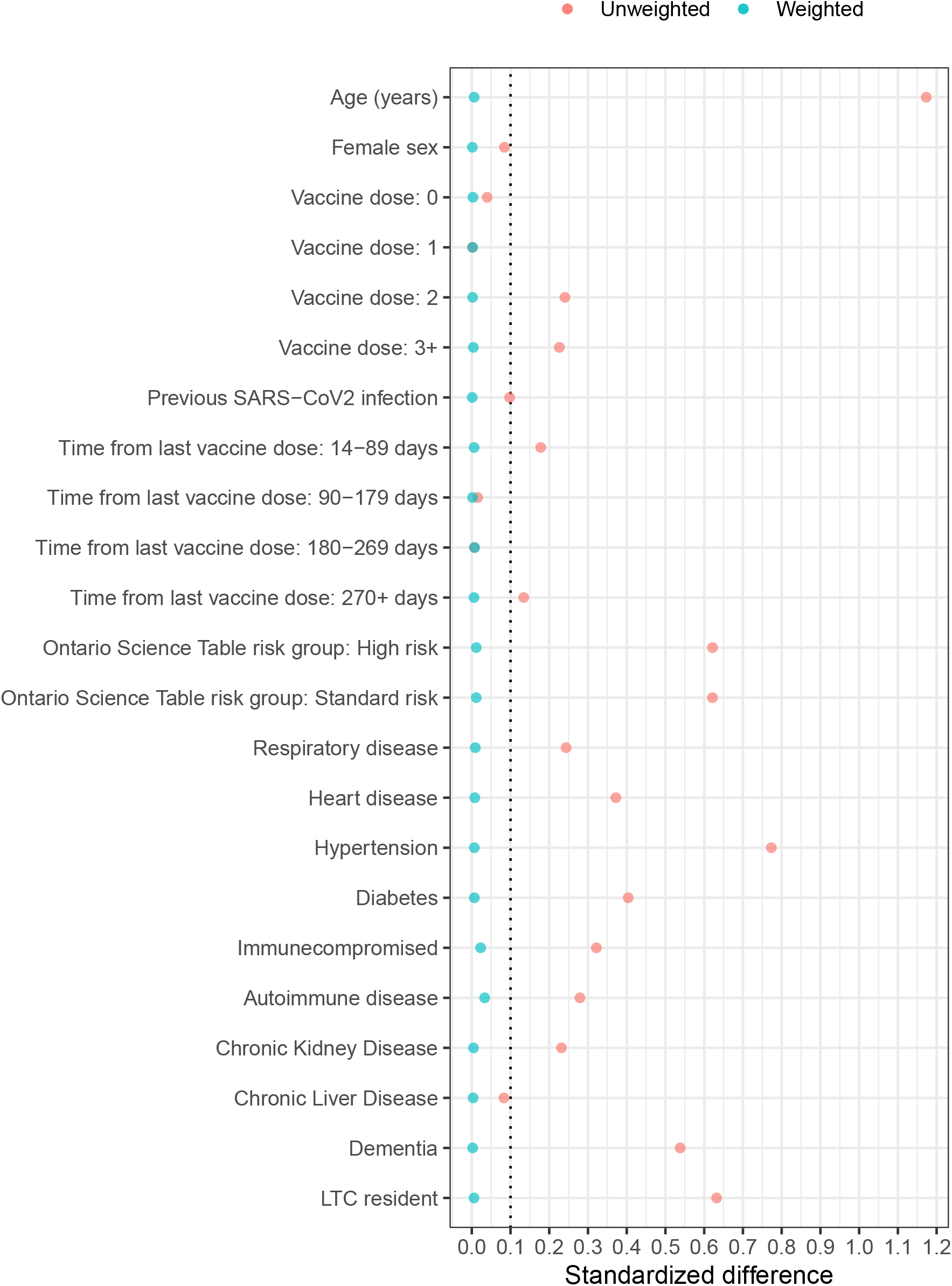
Love plot showing standardized differences (SD) in unweighted cohort and after applying stabilized inverse probability of treatment weighting.

In the weighted primary analysis, nirmatrelvir/ritonavir recipients and unexposed individuals had a 2.1% and 3.7% risk of hospitalization or death, respectively. This equates to a 44% relative reduction in the odds of hospitalization or death within 30 days (wOR 0.56, 95%CI 0.47-0.67, p<0.001) and a 51% relative reduction in the odds of death (wOR 0.49, 95%CI 0.40-0.60, p<0.001). Results were similar in the stratified analyses by age, vaccine status, comorbidities, DDIs, and OST risk status (Table 2 and Figure 3). We observed a possible decrease in effectiveness over time with wOR 0.43 (95%CI, 0.33-0.57) for hospitalization or death between April to June 2022 and wOR 0.67 (95%CI, 0.52-0.86) in July and August 2022 with a similar trend for death alone. (Figure 3).

**Table 2:**
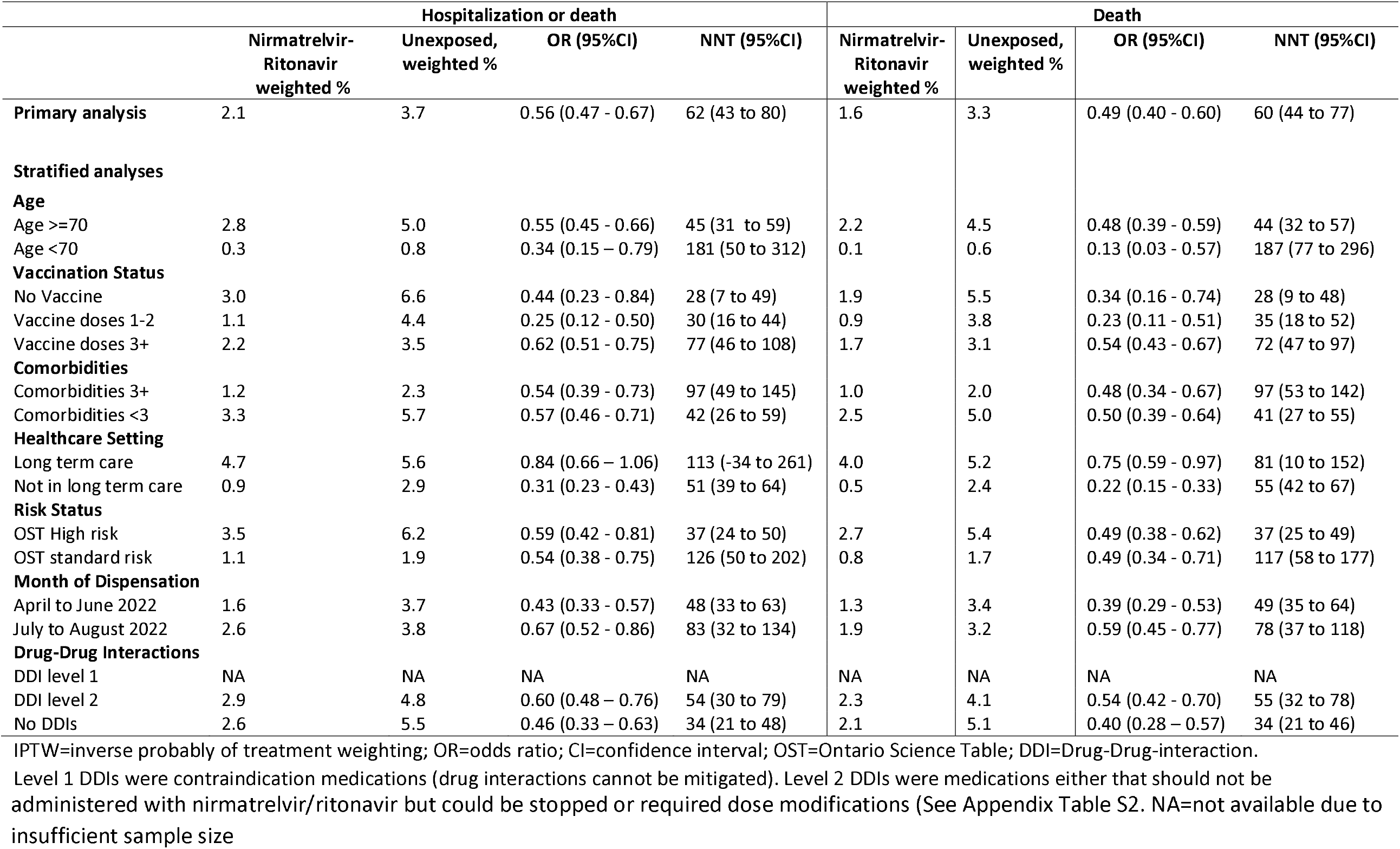
The odds of hospitalization or death, or death, within 30 days of nirmatrelvir/ritonavir receipt (n=8,876), or not (n=168,669), in weighted analysis using IPTW

|  | Hospitalization or death |  |  |  | Death |  |  |  |
| --- | --- | --- | --- | --- | --- | --- | --- | --- |
|  | Nirmatrelvir-Ritonavir<br>weighted % | Unexposed,<br>weighted % | OR (95%CI) | NNT (95%CI) | Nirmatrelvir-Ritonavir<br>weighted % | Unexposed,<br>weighted % | OR (95%CI) | NNT (95%CI) |
| **Primary analysis** | 2.1 | 3.7 | 0.56 (0.47 - 0.67) | 62 (43 to 80) | 1.6 | 3.3 | 0.49 (0.40 - 0.60) | 60 (44 to 77) |
| **Stratified analyses** |  |  |  |  |  |  |  |  |
| **Age** |  |  |  |  |  |  |  |  |
| Age >=70 | 2.8 | 5.0 | 0.55 (0.45 - 0.66) | 45 (31 to 59) | 2.2 | 4.5 | 0.48 (0.39 - 0.59) | 44 (32 to 57) |
| Age <70 | 0.3 | 0.8 | 0.34 (0.15 - 0.79) | 181 (50 to 312) | 0.1 | 0.6 | 0.13 (0.03 - 0.57) | 187 (77 to 296) |
| **Vaccination Status** |  |  |  |  |  |  |  |  |
| No Vaccine | 3.0 | 6.6 | 0.44 (0.23 - 0.84) | 28 (7 to 49) | 1.9 | 5.5 | 0.34 (0.16 - 0.74) | 28 (9 to 48) |
| Vaccine doses 1-2 | 1.1 | 4.4 | 0.25 (0.12 - 0.50) | 30 (16 to 44) | 0.9 | 3.8 | 0.23 (0.11 - 0.51) | 35 (18 to 52) |
| Vaccine doses 3+ | 2.2 | 3.5 | 0.62 (0.51 - 0.75) | 77 (46 to 108) | 1.7 | 3.1 | 0.54 (0.43 - 0.67) | 72 (47 to 97) |
| **Comorbidities** |  |  |  |  |  |  |  |  |
| Comorbidities 3+ | 1.2 | 2.3 | 0.54 (0.39 - 0.73) | 97 (49 to 145) | 1.0 | 2.0 | 0.48 (0.34 - 0.67) | 97 (53 to 142) |
| Comorbidities <3 | 3.3 | 5.7 | 0.57 (0.46 - 0.71) | 42 (26 to 59) | 2.5 | 5.0 | 0.50 (0.39 - 0.64) | 41 (27 to 55) |
| **Healthcare Setting** |  |  |  |  |  |  |  |  |
| Long term care | 4.7 | 5.6 | 0.84 (0.66 - 1.06) | 113 (-34 to 261) | 4.0 | 5.2 | 0.75 (0.59 - 0.97) | 81 (10 to 152) |
| Not in long term care | 0.9 | 2.9 | 0.31 (0.23 - 0.43) | 51 (39 to 64) | 0.5 | 2.4 | 0.22 (0.15 - 0.33) | 55 (42 to 67) |
| **Risk Status** |  |  |  |  |  |  |  |  |
| OST High risk | 3.5 | 6.2 | 0.59 (0.42 - 0.81) | 37 (24 to 50) | 2.7 | 5.4 | 0.49 (0.38 - 0.62) | 37 (25 to 49) |
| OST standard risk | 1.1 | 1.9 | 0.54 (0.38 - 0.75) | 126 (50 to 202) | 0.8 | 1.7 | 0.49 (0.34 - 0.71) | 117 (58 to 177) |
| **Month of Dispensation** |  |  |  |  |  |  |  |  |
| April to June 2022 | 1.6 | 3.7 | 0.43 (0.33 - 0.57) | 48 (33 to 63) | 1.3 | 3.4 | 0.39 (0.29 - 0.53) | 49 (35 to 64) |
| July to August 2022 | 2.6 | 3.8 | 0.67 (0.52 - 0.86) | 83 (32 to 134) | 1.9 | 3.2 | 0.59 (0.45 - 0.77) | 78 (37 to 118) |
| **Drug-Drug Interactions** |  |  |  |  |  |  |  |  |
| DDI level 1 | NA | NA | NA | NA | NA | NA | NA | NA |
| DDI level 2 | 2.9 | 4.8 | 0.60 (0.48 - 0.76) | 54 (30 to 79) | 2.3 | 4.1 | 0.54 (0.42 - 0.70) | 55 (32 to 78) |
| No DDIs | 2.6 | 5.5 | 0.46 (0.33 - 0.63) | 34 (21 to 48) | 2.1 | 5.1 | 0.40 (0.28 - 0.57) | 34 (21 to 46) |

IPTW=inverse probability of treatment weighting; OR=odds ratio; CI=confidence interval; OST=Ontario Science Table; DDI=Drug-Drug-interaction.
Level 1 DDIs were contraindication medications (drug interactions cannot be mitigated). Level 2 DDIs were medications either that should not be

**Figure 3:**
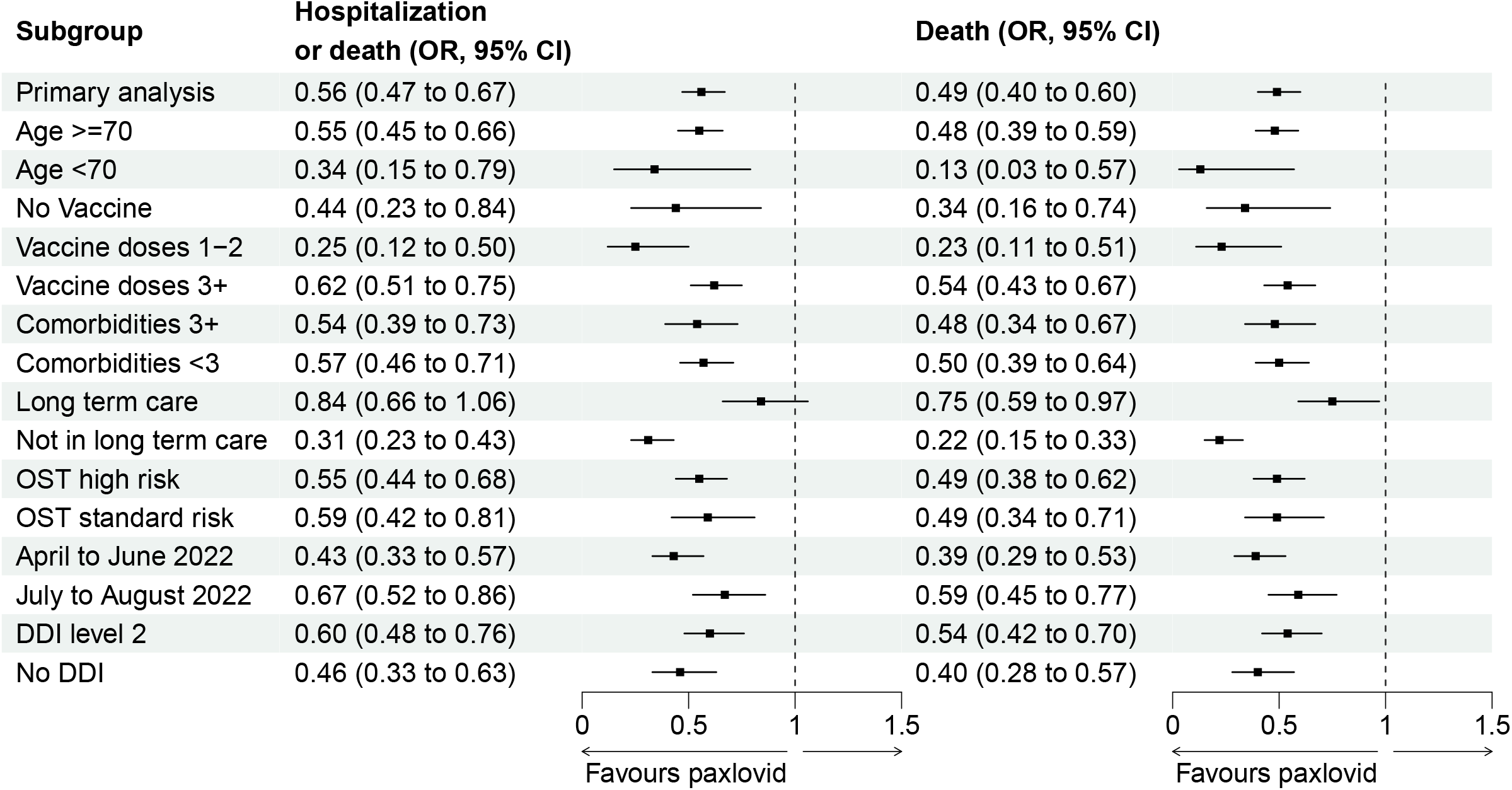
Forest plot of weighted odds ratios with 95%CIs for hospitalization or death from COVID-19, and death, at 30 days for the primary analysis and stratified analyses in patients exposed to nirmatrelvir/ritonavir compared to those unexposed.

Based on this real-world evaluation the NNT was 62 (95%CI 43 to 80) individuals treated with nirmatrelvir/ritonavir to prevent one hospitalization or death from COVID-19. There was substantial variability in absolute risk reductions by strata, with NNT ranging from 28 (95%CI 7 to 49) for unvaccinated individuals to 180 (95%CI 50 to 312) for those <70 years of age (Table 2 and Figure S2).

## Interpretation

The use of nirmatrelvir/ritonavir in Ontario, Canada between April and August 2022 was associated with a 44% relative reduction in the odds of hospitalization or death from COVID-19. Findings were consistent across most strata of age, DDIs, vaccination status, and comorbidities. The largest benefits, both in terms of absolute and relative risk reduction were observed in undervaccinated or unvaccinated patients and individuals 70 years of age or older.

The EPIC-HR study is the only published randomized controlled trial on nirmatrelvir/ritonavir which demonstrated a relative risk reduction of 89% and an absolute risk reduction of 6% (NNT=16) in severe COVID-19. However, this study was conducted prior to the Omicron variant and excluded vaccinated patients and those with DDIs.(2) In a subsequent yet unpublished study in lower risk patients, EPIC-SR, a company press release reported a non-significant 57% relative reduction in hospitalizations and death.(17) In our population who received nirmatrelvir/ritonavir, approximately 5% were unvaccinated, the mean age was 74 years of age, 67% of those 70 years or older had potentially significant DDIs, and 32% resided in long term care, thus representing a very different patient population compared to the clinical trial. After adjustment for substantial confounding and addressing potential immortal time bias through the study design, we observed a significant clinical benefit to nirmatrelvir/ritonavir use, albeit attenuated in both relative and absolute risk reduction compared to the EPIC-HR trial. This is likely due to differences in the patient population, underlying immunity in the population, and decreased severity between circulating variants.

A recent meta-analysis attempted to summarize the safety and effectiveness of nirmatrelvir/ritonavir from published trials and observational studies and reported an OR of death and hospitalization of 0.22 (95%CI, 0.11-0.45) with no safety concerns.(18) A cohort study from Israel identified an adjusted hazard ratio for severe COVID-19 in patients 65 years of age or older of 0.27 (95%CI: 0.15-0.49), but only 0.74 (95%CI: 0.35-1.58) in those 40-64 years of age. Their findings were similar when further stratified by previous immunity to SARS-CoV-2. This study was also a real world evaluation with similar patient characteristics to ours although notably the Israeli cohort excluded patients with DDIs.(3)

Our study, in conjunction with previous clinical trials and observational research, supports the effectiveness of nirmatrelvir/ritonavir at reducing hospitalization and death from COVID-19. Risk factors for severe disease include older age as the single most important risk factor, as well as obesity, the number of comorbidities, and time from previous vaccination. Vaccination and immunity from prior infection are significantly protective.(19) The OST delineated high and standard risk population in Ontario to prioritize limited initial medication supplies. Our study identified that both groups have similar relative risk reductions in severe COVID-19, however there are substantial differences in the absolute risk reductions which have implications for cost-effectiveness evaluations for a publically funded therapeutic. The estimated NNT to prevent one hospitalization or death from COVID-19 for high and standard OST risk criteria were 37 and 126, respectively.

We found that there were few patients older than 70 years of age with level 1 (contraindicated) DDIs who received nirmatrelvir/ritonavir. Two-thirds of our cohort 70 years of age or older had potential level 2 (caution) DDIs reflecting the numerous medications that have clinically significant DDI with ritonovir. Based on the data sources used in this study, we were unable to confirm whether patients were actually taking interacting medications concurrently or to determine if any potential DDI were appropriately mitigated at the time of nirmatrelvir/ritonavir prescribing which may have impacted our estimation of DDIs. Our results are encouraging that patients with level 2 DDIs can be effectively treated with nirmatrelvir/ritonavir and the importance of prescribers and pharmacists evaluating for, and mitigating DDIs.

It is notable that we observed a potential decrease in effectiveness in the latter two months of our study. This finding may be due to chance, changing severity over time, or potentially antiviral resistance. Resistance in SARS-CoV-2 to nirmatrelvir/ritonavir is possible,(21) however no resistance has been demonstrated as of yet among Omicron and its subvariants.(22) The latter two months of the study were characterized by growth of the BA.5 omicron subvariant and notably the rate of the outcome in the control group appears decreased. Further observation of therapeutic effectiveness for current and future variants is essential.

This study has some notable limitations. The cohort was limited to those testing positive for SARS-CoV-2 by PCR and does not include those who may have tested positive only by rapid antigen tests. We therefore could not include all patients who received nirmatrelvir/ritonavir which may limit the generalizability of the findings. Nirmatrelvir/ritonavir was publically funded through ODB for all Ontario residents, however not all dispenses in Ontario are captured in ODB. We could ascertain that approximately 85% of nirmatrelvir/ritonavir courses are in ODB and we excluded patients tested from any of the 27 COVID-19 assessment centres where nirmatrelvir/ritonavir is dispensed without an ODB claim to limit the risk of misclassification bias. Our database indicates that the medication was dispensed but we could not assess adherence. Observational cohort studies have a risk of immortal time bias which we attempted to mitigate through imputing theoretical dispensing dates to the unexposed group. By imputing index dates that mirror the exposed groups’ distribution we removed unexposed individuals from the cohort who may have experienced the outcome prior to having the opportunity to receive nirmatrelvir/ritonavir; thereby mitigating immortal time bias. However, some residual bias is possible. There is significant confounding in the underlying data as nirmatrelvir/ritonavir is recommended and limited to patients who were at higher risk of the outcome. We demonstrated that the IPTW method successfully balanced the groups by all evaluated covariates but residual confounding is possible.

In conclusion, in this population-level real world evaluation of nirmatrelvir/ritonavir we observed a significant risk reduction in hospitalization and death from COVID-19 supporting ongoing use of this antiviral to treat patients with mild illness who are at risk of severe COVID-19. The relative effectiveness was similar across the evaluated strata and risk groups with substantial variation in the absolute risk reduction, or NNT, with implications for cost-effectiveness evaluations. Ongoing evaluation to monitor effectiveness in the population with new circulating variants is critical to inform optimal use over time.

## Data Availability

The dataset from this study is held securely in coded form at ICES. While legal data sharing agreements between ICES and data providers (e.g., healthcare organizations and government) prohibit ICES from making the dataset publicly available, access may be granted to those who meet pre-specified criteria for confidential access, available at www.ices.on.ca/DAS. The full dataset creation plan and underlying analytic code are available from the authors upon request, understanding that the computer programs may rely upon coding templates or macros that are unique to ICES and are therefore either inaccessible or may require modification.

## Acknowledgments

This study was supported by ICES, which is funded by an annual grant from the Ontario Ministry of Health (MOH) and the Ministry of Long-Term Care (MLTC). This study was also funded by Public Health Ontario. The analyses, conclusions, opinions and statements expressed herein are solely those of the authors and do not reflect those of the funding or data sources; no endorsement is intended or should be inferred.

Parts of this material are based on data and/or information compiled and provided by CIHI. However, the analyses, conclusions, opinions and statements expressed in the material are those of the author(s), and not necessarily those of CIHI.

We thank IQVIA Solutions Canada Inc. for use of their Drug Information File.

