## Appendix for "Real-world effectiveness of nirmatrelvir/ritonavir use for COVID-19: A population-based cohort study in Ontario, Canada"

Table S1: ICES Covariate definitions

| Variable | Definition |
| --- | --- |
| Age | Age was determined from the Registered Persons Database. |
| Sex | Sex was determined from the Registered Persons Database. |
| Chronic heart disease | Individuals were defined as having “chronic heart disease” if they had congestive heart failure (CHF), ischemic heart disease, or atrial fibrillation. The definitions for these conditions are as follows:  CHF:^2^  An ICES-derived CHF database was used to identify patients with CHF, based on 1 NACRS, DAD, SDS, or OHIP claim and a second claim (from either) in 1 year. The CHF database is limited to those aged 40 years or older.  OHIP: 428  DAD, SDS: ICD-9: 428, ICD-10: I500, I501, I509  Cardiac ischemic disease:^4^  Any comorbidity in the past 5 years (DAD, any diagnosis field) or history of procedure in past 20 years (DAD, SDS), of the following:    Comorbidity (DAD, any diagnosis in the past 5 years):  Angina: ICD-10: I20    Chronic Ischemic Heart Disease: ICD-10: I25;  Myocardial infarction: ICD-10: I21, I22  Procedure (DAD & SDS):  Coronary Artery Bypass Grafting:  CCI procedure codes: 1IJ76  CCP procedure codes: 481    Percutaneous Coronary Intervention:  CCI procedure codes: 1IJ50, 1IJ54, 1IJ57GQ  CCP procedure codes: 4802, 4803  Atrial fibrillation:^5^  Individuals with 1 hospitalization or 4 MD visits within a year in the past 5 years with the following codes:  ICD-9: 427.31, 427.32  ICD-10: I48  OHIP dxcode: 427 |
| Chronic respiratory disease | Asthma:^6^  An ICES-specific asthma database was used to identify patients with  asthma, based on 2 or more ambulatory care visits and/or 1 or more  hospitalizations.  OHIP  OHIP diagnostic code: 493  DAD  ICD-9 diagnostic code: 493  ICD-10 diagnostic codes: J45, J46  Chronic obstructive pulmonary disease (COPD):^7^  An ICES-specific COPD database was used to identify patients with  COPD, based on 1 or more ambulatory care visits and/or 1 or more  hospitalizations. The algorithm to identify COPD patients was only  validated in those aged 35 years or older.  OHIP  OHIP diagnostic codes: 491, 492, 496  DAD  ICD-9 diagnostic codes: 491, 492, 496  ICD-10 diagnostic codes: J41, J42, J43, J44 |
| Hypertension | An ICES-specific hypertension database was used to identify patients with hypertension, based on 1 or more DAD diagnoses or 2 or more OHIP diagnoses in a two-year period; or 1 OHIP diagnosis followed by an OHIP/DAD diagnosis within two years.^8^  DAD, SDS: ICD-9: 401, 402, 403, 404, 405; ICD-10: I10, I11, I12, I13, I15  OHIP diagnostic codes: 401, 402, 403, 404, or 405 |
| Diabetes | An ICES-specific diabetes database was used to identify patients with diabetes, based on 2 OHIP diagnostic codes or 1 OHIP service code or 1 DAD admission within 2 years.^9^  DAD, SDS: ICD-9: 250; ICD-10: E10, E11, E13, E14  OHIP: 250  OHIP service codes: Q040, K029, K030, K045, K046 |
| Immunocompromised (HIV, transplant, immunosuppressive therapy) | HIV:^10^  An ICES-specific HIV database was used to identify patients with HIV, based on 3 physician claims in 3 years with OHIP diagnostic codes: 042, 043 or 044  Solid organ transplant recipients:  CORRLINK is an ICES-specific database which links CORR (Canadian Organ Replacement Register) and DAD data. This database only includes patients that have received an organ transplant and does not include dialysis patients.   - For transplants before December 31, 2019: individuals are a transplant recipient if they have a treatment code of 171, where the treatment was before the index date - For transplants on/after January 1, 2020: Identify ICD-10 codes, CCI procedure codes, and OHIP feecodes from DAD, NACRS, and OHIP (codes available upon request)     Allogenic/autologous bone marrow transplant recipients:  We identified those who had a history of allogenic bone marrow transplant before the index date using the following combination of diagnostic codes:  DAD:   - CCP procedure codes = 53.0 - CCI procedure codes = 1WY19, 1LZ19HHU7, 1LZ19HHU8   OHIP:   - Feecode = Z426     Other immune disorders:  Individuals were identified as having disorders of the immune system based on health care encounters recorded in DAD, SDS, NACRS, and OHIP in the 2-years prior to index using Expanded Diagnostic Clusters from the Johns Hopkins ACGⓇ System Version 10.^11^  Any hospitalization (any diagnosis field) with the following codes:   - Sickle-cell disease (ICD-10 D57.0 – D57.2; D57.8 OR ICD-9 282.6); - Other immune system disorders (ICD-9 273.2, 279.0, 279.1, 279.2, 279.3, 279.8, 279.9, 289.8; ICD-10 D80, D81, D82, D83, D84, D89; OHIP dxcode 279) - Immunosuppressive therapy (>30 days (total days supplied) of oral corticosteroid in the 6 months before index date; receipt of other immunocompromising drug, including antineoplastics, in the 6 months before index date)   Active cancer:   - Any of the following treatments in the past 6 months: cancer surgery (codes available upon request), radiation (if the ICD-10 code listed was Z510 in NACRS), chemotherapy (if the ICD-10 code listed was Z511 or Z512 and any evidence of cancer diagnosis in the Ontario Cancer Registry (OCR) prior to the last treatment date) - If not any of the above, individuals still classified as having cancer if they had a cancer diagnosis in OCR in the year before the index date |
| Autoimmune disease | Individuals considered to have autoimmune disease if they had any of the following:   - Rheumatoid arthritis (identified in the Ontario Rheumatoid Arthritis Database)^12,13^ - Inflammatory bowel disease (identified in the Ontario Crohn’s and Colitis Cohort)^14,15^ - Psoriasis/psoriatic arthritis^16^   - Psoriasis: 1 hospitalization or 3 physician billings prior to index date. DAD: ICD-9: 696.1, 696.8; ICD-10: L40.0, L40.1, L40.2, L40.3, L40.4, L40.8, L40.9. OHIP: dxcode = 696   - Psoriatic arthritis: 1 hospitalization or: (3 physician billings for psoriatic arthritis + 1 billing for psoriasis [696]). DAD: ICD-9: 696.0; ICD-10: L40.5, M07.0, M07.1, M07.2, M07.3, M09.0. OHIP: dxcode = 721 (at least one of these billings must be billed by a rheumatologist). - Multiple sclerosis^17^   - Individuals with one hospitalization or 5 physician billings over 2 years. DAD: ICD-9: 340; ICD-10: G35. OHIP: dxcode = 340. |
| Chronic kidney disease (CKD) | We defined this variable as having a CKD diagnosis code in DAD, NACRS, OHIP in the past 5 years, or: at least 1 dialysis code in each of the 3 months prior to index.^18^  OHIP: 403, 585  ICD-10: E102, E112, E132, E142, I12, I13, N08, N18, N19 |
|  | Patients who were on chronic dialysis in the year before index date, identified as those with at least 2 of any of the following codes in OHIP, DAD, or SDS separated by at least 90 days, but less than 150 days:^19^  OHIP service codes: R849, G323, G325, G326, G860, G862, G865 G863, G866, G330, G331, G332, G333, G861, G082, G083, G085, G090, G091, G092, G093, G094, G095, G096, G294, G295, G864, H540, H740  DAD, SDS:  CCI procedure codes: 5195, 6698  CCP procedure code: 1PZ21 |
| Dementia | Dementia (ICES cohort) definition:  1 hospitalization for dementia and/or 3 ambulatory visits for dementia, each separated by at least 30 days, within 2 years and/or 1 prescription from ODB.^21^  OHIP: 290, 331  DAD, SDS:  ICD-9: 0461, 290.0, 290.1, 290.2, 290.3, 290.4, 294, 331.0, 331.1, 331.5  ICD-10: F00, F01, F02, F03, G30  ODB:  1 prescription for a cholinesterase inhibitor |
| Advanced liver disease (Cirrhosis or Decompensated Cirrhosis) | Defined using the Cirrhosis Algorithm 9 [from ^20^]:  Two or more physician visits (diagnosis code 571), or  one or more hospital diagnosis of cirrhosis, using the following diagnostic codes:  ICD-9 : 456.1, 571.2, 571.5  ICD-10: I85.9, I98.2, K70.3,K71.7, K74.6  Defined using the Decompensated Cirrhosis Algorithm 5 (from ^20^):  One or more physician visits with diagnosis code 571 and (one or more hospital diagnosis or one or more procedure), using the following diagnostic codes:  ICD-9: 456.0, 456.2, 572.2, 572.3, 572.4, 782.4, 789.5l; ICD-10: I85.0, I86.4, I98.20, I98.3, K721, K729, K76.6, K76.7, R17, R18  CCI: 1.NA.13.BA-FA, 1.NA.13.BA-X7, 1.NA.13.BA-BD, 1.KQ.76GP-NR, 1.OT.52.HA  CCP: 1006, 6691  OHIP: J057, Z591 |

Table S2: Ontario Ministry of Health criteria for nirmatrelvir/ritonavir

- Immunocompromised (have an immune system that is weakened by a health condition or medications)
- 70 and older
- 60 and older with less than three vaccine doses
- 18 and older with less than three vaccine doses and at least one of the following risk conditions:
  - obesity
  - diabetes
  - heart disease
  - hypertension
  - congestive heart failure
  - chronic respiratory disease (including cystic fibrosis)
  - cerebral palsy
  - intellectual or developmental disability
  - sickle cell disease
  - moderate or severe kidney disease
  - moderate or severe liver disease
  - pregnancy

Figure S1: Time-To-Dispense: Days from positive SARS-CoV-2 test to nirmatrelvir/ritonovir dispensing

Percent

| Day from test for SARS-CoV-2 | Cases | Controls (imputed) |
| --- | --- | --- |
| 0 | 37.0914 | 37.0106 |
| 1 | 30.9646 | 31.1772 |
| 2 | 17.9023 | 17.7443 |
| 3 | 8.65505 | 8.71891 |
| 4 | 3.70117 | 3.68141 |
| 5 | 1.68546 | 1.66757 |

Table S2: Drug-drug interactions with nirmatrelvir/ritonavir

| **Drug Class (AHFS Classification)** | **Drug Name** | **DDI Severity, 1=contraindicated, 2=mitigation required** |
| --- | --- | --- |
| **08:00 Anti-infective Agents** |  |  |
| 08:00 Anti-infective Agents | Rifabutin | 2 |
| 08:00 Anti-infective Agents | Rifampin | 1 |
| 08:00 Anti-infective Agents | Rifapentine | 1 |
| 08:00 Anti-infective Agents | Glecaprevir/Pibrentasvir | 2 |
| 08:00 Anti-infective Agents | Quinine | 2 |
| **10:00 Antineoplastic Agents** |  |  |
| 10:00 Antineoplastic Agents | Abemaciclib | 2 |
| 10:00 Antineoplastic Agents | Apalutamide | 1 |
| 10:00 Antineoplastic Agents | Bosutinib | 2 |
| 10:00 Antineoplastic Agents | Ceritinib | 2 |
| 10:00 Antineoplastic Agents | Cobimetinib | 2 |
| 10:00 Antineoplastic Agents | Dabrafenib | 1 |
| 10:00 Antineoplastic Agents | Dasatinib | 2 |
| 10:00 Antineoplastic Agents | Encorafenib | 2 |
| 10:00 Antineoplastic Agents | Enzalutamide | 1 |
| 10:00 Antineoplastic Agents | Ibrutinib | 2 |
| 10:00 Antineoplastic Agents | Lorlatinib | 1 |
| 10:00 Antineoplastic Agents | Mitotane | 1 |
| 10:00 Antineoplastic Agents | Neratinib | 2 |
| 10:00 Antineoplastic Agents | Nilotinib | 2 |
| 10:00 Antineoplastic Agents | Sonidegib | 1 |
| 10:00 Antineoplastic Agents | Tepotinib | 1 |
| 10:00 Antineoplastic Agents | Venetoclax | 1 |
| 10:00 Antineoplastic Agents | vinBLAStine | 2 |
| 10:00 Antineoplastic Agents | vinCRIStine | 2 |
| **12:12 Sympathomimetic Agents** |  |  |
| 12:12 Sympathomimetic Agents | Salmeterol | 2 |
| 12:12 Sympathomimetic Agents | Alfuzosin | 2 |
| 12:12 Sympathomimetic Agents | Silodosin | 2 |
| 12:12 Sympathomimetic Agents | Tamsulosin | 2 |
| **20:00 Blood Formation, Coagulation and Thrombosis Agents** | | |
| 20:00 Blood Formation, Coagulation and Thrombosis Agents | Warfarin | 2 |
| 20:00 Blood Formation, Coagulation and Thrombosis Agents | Dabigatran | 2 |
| 20:00 Blood Formation, Coagulation and Thrombosis Agents | Apixaban | 2 |
| 20:00 Blood Formation, Coagulation and Thrombosis Agents | Edoxaban Tosylate | 2 |
| 20:00 Blood Formation, Coagulation and Thrombosis Agents | Rivaroxaban | 2 |
| 20:00 Blood Formation, Coagulation and Thrombosis Agents | Clopidogrel | 2 |
| 20:00 Blood Formation, Coagulation and Thrombosis Agents | Ticagrelor | 2 |
| 20:00 Blood Formation, Coagulation and Thrombosis Agents | Fostamatinib | 2 |
| **24:00 Cardiovascular Drugs** |  |  |
| 24:00 Cardiovascular Drugs | quiNIDine | 1 |
| 24:00 Cardiovascular Drugs | Phenytoin | 1 |
| 24:00 Cardiovascular Drugs | Flecainide | 1 |
| 24:00 Cardiovascular Drugs | Propafenone | 1 |
| 24:00 Cardiovascular Drugs | Amiodarone | 1 |
| 24:00 Cardiovascular Drugs | Dofetilide | 2 |
| 24:00 Cardiovascular Drugs | Dronedarone | 1 |
| 24:00 Cardiovascular Drugs | Digoxin | 2 |
| 24:00 Cardiovascular Drugs | Ranolazine | 1 |
| 24:00 Cardiovascular Drugs | Atorvastatin | 2 |
| 24:00 Cardiovascular Drugs | Lovastatin | 2 |
| 24:00 Cardiovascular Drugs | Rosuvastatin | 2 |
| 24:00 Cardiovascular Drugs | Simvastatin | 2 |
| 24:00 Cardiovascular Drugs | Lomitapide | 2 |
| 24:00 Cardiovascular Drugs | Sildenafil | 2 |
| 24:00 Cardiovascular Drugs | Tadalafil | 2 |
| 24:00 Cardiovascular Drugs | Vardenafil | 2 |
| 24:00 Cardiovascular Drugs | Bosentan | 1 |
| **24:28 Calcium-Channel Blocking Agents** | |  |
| 24:28 Calcium-Channel Blocking Agents | amLODIPine | 2 |
| 24:28 Calcium-Channel Blocking Agents | Felodipine | 2 |
| 24:28 Calcium-Channel Blocking Agents | NIFEdipine | 2 |
| 24:28 Calcium-Channel Blocking Agents | dilTIAZem | 2 |
| 24:28 Calcium-Channel Blocking Agents | Verapamil | 2 |
| **28:00 Central Nervous System Drugs** | |  |
| 28:00 Central Nervous System Drugs | fentaNYL | 1 |
| 28:00 Central Nervous System Drugs | HYDROcodone | 2 |
| 28:00 Central Nervous System Drugs | Meperidine | 2 |
| 28:00 Central Nervous System Drugs | oxyCODONE | 2 |
| 28:00 Central Nervous System Drugs | traMADol | 2 |
| 28:00 Central Nervous System Drugs | PHENobarbital | 1 |
| 28:00 Central Nervous System Drugs | Primidone | 1 |
| 28:00 Central Nervous System Drugs | Phenytoin | 1 |
| 28:00 Central Nervous System Drugs | carBAMazepine | 1 |
| 28:00 Central Nervous System Drugs | Eslicarbazepine | 1 |
| 28:00 Central Nervous System Drugs | Oxcarbazepine | 1 |
| 28:00 Central Nervous System Drugs | ARIPiprazole | 2 |
| 28:00 Central Nervous System Drugs | Brexpiprazole | 2 |
| 28:00 Central Nervous System Drugs | cloZAPine | 1 |
| 28:00 Central Nervous System Drugs | Lurasidone | 1 |
| 28:00 Central Nervous System Drugs | QUEtiapine | 2 |
| 28:00 Central Nervous System Drugs | risperiDONE | 2 |
| 28:00 Central Nervous System Drugs | Ziprasidone | 2 |
| 28:00 Central Nervous System Drugs | Pimozide | 1 |
| 28:00 Central Nervous System Drugs | Modafinil | 2 |
| 28:00 Central Nervous System Drugs | ALPRAZolam | 2 |
| 28:00 Central Nervous System Drugs | Clorazepate | 2 |
| 28:00 Central Nervous System Drugs | diazePAM | 2 |
| 28:00 Central Nervous System Drugs | Flurazepam | 2 |
| 28:00 Central Nervous System Drugs | Midazolam | 2 |
| 28:00 Central Nervous System Drugs | Triazolam | 2 |
| 28:00 Central Nervous System Drugs | clonazePAM | 2 |
| 28:00 Central Nervous System Drugs | Nitrazepam | 2 |
| 28:00 Central Nervous System Drugs | busPIRone | 2 |
| 28:00 Central Nervous System Drugs | Zolpidem | 2 |
| 28:00 Central Nervous System Drugs | Zopiclone | 2 |
| 28:00 Central Nervous System Drugs | Dihydroergotamine Mesylate | 2 |
| 28:00 Central Nervous System Drugs | Ergotamine | 2 |
| **56:00 Gastrointestinal Drugs** |  |  |
| 56:00 Gastrointestinal Drugs | Cisapride | 2 |
| **68:00 Hormones and Synthetic Substitutes** | |  |
| 68:00 Hormones and Synthetic Substitutes | Dexamethasone | 2 |
| 68:00 Hormones and Synthetic Substitutes | Elagolix | 2 |
| **76:00 Oxytocics** |  |  |
| 76:00 Oxytocics | Ergonavine/Methylergonovine | 2 |
| **92:00 Miscellaneous Therapeutic Agents** | |  |
| 92:00 Miscellaneous Therapeutic Agents | Colchicine | 2 |
| 92:00 Miscellaneous Therapeutic Agents | cycloSPORINE | 2 |
| 92:00 Miscellaneous Therapeutic Agents | Sirolimus | 2 |
| 92:00 Miscellaneous Therapeutic Agents | Tacrolimus | 2 |
| 92:00 Miscellaneous Therapeutic Agents | Everolimus | 2 |
| 92:00 Miscellaneous Therapeutic Agents | Tacrolimus | 2 |
| **Other** |  |  |
| Other | St. John's Wort | 1 |
| **Source**: Ontario COVID-19 Drugs and Biologics Clinical Practice Guidelines Working Group, University of Waterloo School of Pharmacy. Nirmatrelvir/Ritonavir (Paxlovid): What prescribers and pharmacists need to know. Ontario COVID-19 Science Advisory Table. 2022;3(58). https://doi.org/10.47326/ocsat.2022.03.58.1.0 | | |

Table S3: Pre-weighted variable numbers for stratification

| **variable** | **Total n=177,545** | **Unexposed n=168,669** | **Exposed n=8,876** |
| --- | --- | --- | --- |
| **Age <70y** | 132,090 (74.4%) | 129,647 (76.9%) | 2,443 (27.5%) |
| **Age 70+y** | 45,455 (25.6%) | 39,022 (23.1%) | 6,433 (72.5%) |
| **>0 vaccine doses** | 166,644 (93.9%) | 158,235 (93.8%) | 8,409 (94.7%) |
| **0 vaccine doses** | 10,901 (6.1%) | 10,434 (6.2%) | 467 (5.3%) |
| **<1 or 3+ vaccine doses** | 146,331 (82.4%) | 138,340 (82.0%) | 7,991 (90.0%) |
| **1-2 vaccine doses** | 31,214 (17.6%) | 30,329 (18.0%) | 885 (10.0%) |
| **<3 vaccine doses** | 42,115 (23.7%) | 40,763 (24.2%) | 1,352 (15.2%) |
| **3+ vaccine doses** | 135,430 (76.3%) | 127,906 (75.8%) | 7,524 (84.8%) |
| **Comorbidities <3** | 146,852 (82.7%) | 141,781 (84.1%) | 5,071 (57.1%) |
| **Comorbidities 3+** | 30,693 (17.3%) | 26,888 (15.9%) | 3,805 (42.9%) |
| **OST Standard risk** | 148,326 (83.5%) | 143,170 (84.9%) | 5,156 (58.1%) |
| **OST High risk** | 29,219 (16.5%) | 25,499 (15.1%) | 3,720 (41.9%) |
| **April to June 2022** | 109,961 (61.9%) | 105,755 (62.7%) | 4,206 (47.4%) |
| **July to August 2022** | 67,584 (38.1%) | 62,914 (37.3%) | 4,670 (52.6%) |
| **No DDI or level 2*** | 44,616 (98.2%) | 38,217 (97.9%) | 6,399 (99.5%) |
| **DDI level 1*** | 839 (1.8%) | 805 (2.1%) | 34 (0.5%) |
| **No DDI or level 1*** | 16,856 (37.1%) | 14,680 (37.6%) | 2,176 (33.8%) |
| **DDI level 2*** | 28,599 (62.9%) | 24,342 (62.4%) | 4,257 (66.2%) |
| **DDI level 1 or 2*** | 29,438 (64.8%) | 25,147 (64.4%) | 4,291 (66.7%) |
| **No DDI*** | 16,017 (35.2%) | 13,875 (35.6%) | 2,142 (33.3%) |
| **Not a LTC resident** | 161,944 (91.2%) | 155,863 (92.4%) | 6,081 (68.5%) |
| **LTC resident** | 15,601 (8.8%) | 12,806 (7.6%) | 2,795 (31.5%) |

*Limited to those 70 years of age or older

Figure S2: Forest plot of number needed to treat (NNT) with 95%CIs for hospitalization or death and death at 30 days between those exposed to nirmatrelvir/ritonavir and unexposed groups using weighted logistic regression models with IPTW.

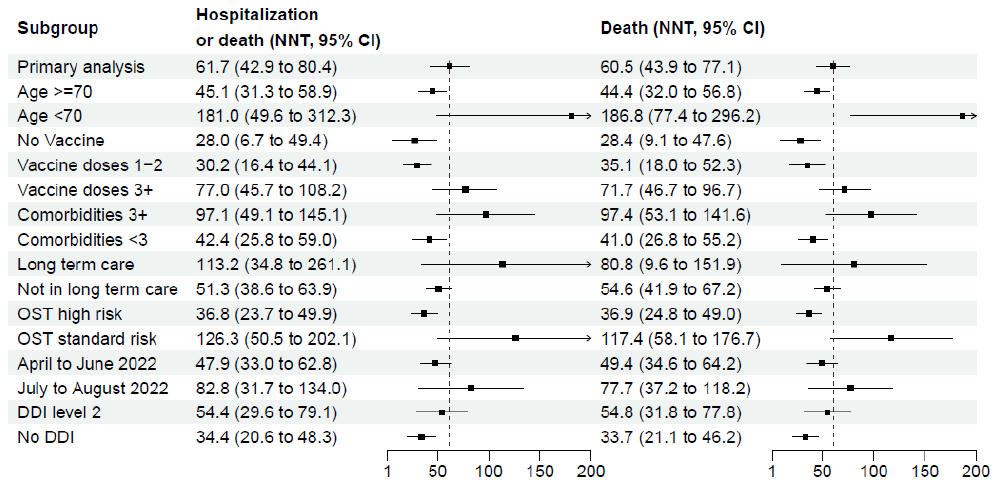
